# Early White Matter Microstructure Alterations in Infants with Down Syndrome

**DOI:** 10.1101/2025.02.26.25322913

**Authors:** Omar Azrak, Dea Garic, Aleeshah Nasir, Meghan R. Swanson, Rebecca L. Grzadzinski, Khalid Al-Ali, Mark D. Shen, Jessica B. Girault, Tanya St. John, Juhi Pandey, Lonnie Zwaigenbaum, Annette M. Estes, Jason J. Wolff, Stephen R. Dager, Robert T. Schultz, Alan C. Evans, Jed T. Elison, Essa Yacoub, Sun Hyung Kim, Robert C. McKinstry, Guido Gerig, John R. Pruett, Joseph Piven, Kelly N. Botteron, Heather Hazlett, Natasha Marrus, Martin A. Styner, Infant Brain Imaging Study (IBIS) Network

## Abstract

**Importance:** Down syndrome, resulting from trisomy 21, is the most prevalent chromosomal disorder and a leading cause of intellectual disability. Despite its significant impact on brain development, research on the white matter microstructure in infants with Down syndrome remains limited.

**Objective:** To investigate early white matter microstructure in infants with Down syndrome using diffusion tensor imaging (DTI) and neurite orientation dispersion and density imaging (NODDI).

**Design:** Infants were recruited and scanned between March 2019 and May 2024 as participants in prospective studies conducted by the Infant Brain Imaging Study (IBIS) Network. Data were analyzed in October 2024.

**Setting:** Data collection occurred at five research centers in Minnesota, Missouri, North Carolina, Pennsylvania, and Washington.

**Participants:** Down syndrome and control infants were scanned at 6 months of age. Control infants had no Down syndrome diagnosis and either had a typically developing older sibling or, if they had an older sibling with autism, were confirmed not to meet clinical best estimate criteria for an autism diagnosis.

**Exposure:** Diagnosis of Down syndrome.

**Main Outcomes and Measures:** The outcome of interest was white matter microstructure quantified using DTI and NODDI measures.

**Results:** A total of 49 Down syndrome (28 [57.14%] female) and 37 control (18 [48.65%] female) infants were included. Infants with Down syndrome showed significant reductions in fractional anisotropy and neurite density index across multiple association tracts, particularly in the inferior fronto-occipital fasciculus and superior longitudinal fasciculus II, consistent with reduced structural integrity and neurite density. These tracts also demonstrated increased radial diffusivity, suggesting delayed myelination. The inferior fronto-occipital fasciculus and uncinate fasciculus exhibited increased neurite dispersion and fanning in Down syndrome infants, reflected by elevated orientation dispersion index. Notably, the optic tracts in Down syndrome infants exhibited a distinct pattern of elevated fractional anisotropy and axial diffusivity, and lower radial diffusivity and orientation dispersion index, suggesting an early maturation of these pathways.

**Conclusions and Relevance:** This first characterization of white matter microstructure in Down syndrome infants reveals widespread white matter developmental delays. These findings provide new insights into the early neurodevelopment of Down syndrome and may inform early therapeutic interventions.

## Introduction

Down syndrome (DS), caused by trisomy 21, is the most common genetic cause of intellectual disability, affecting approximately 1 in 600 newborns.^1,2^ It is a lifelong neurodevelopmental disorder characterized by heterogeneous presentation, susceptibility to regression in childhood, and increased incidence of Alzheimer’s disease (AlzD) in adulthood.^3–5^ Neuroimaging research in DS has primarily focused on older children and adults, consistently revealing volumetric brain reductions and white matter (WM) integrity alterations that contribute to cognitive and functional challenges.^6–9^ While previous studies in infants with DS have examined brain volume,^10–12^ no research has investigated WM microstructure at this early stage.

Infancy is a critical period of brain development, particularly in WM maturation, marked by rapid myelination, heightened plasticity, and the establishment of neural pathways.^13,14^ Understanding neurodevelopmental differences at infancy can (a) provide insight into how atypical neurodevelopmental trajectories emerge, (b) establish a foundation for longitudinal studies to determine how these trajectories evolve and whether they predict later behavioral and functional outcomes, and (c) identify optimal windows for mechanistically-informed interventions aimed at improving long-term cognitive and adaptive functioning in DS.^15,16^

Diffusion MRI, predominantly diffusion tensor imaging (DTI), has been instrumental in identifying WM abnormalities such as changes in fiber integrity and microstructural disruptions associated with demyelination and axonal damage.^17–19^ DTI assumes a single dominant fiber orientation per voxel, making it difficult to interpret in regions with intersecting fibers.^20^ To address this challenge, we employ Neurite Orientation Dispersion and Density Imaging (NODDI) to obtain a more nuanced view of WM microstructure.

In children and young adults, WM abnormalities have been identified in key tracts, including the inferior fronto-occipital fasciculus (IFOF), inferior longitudinal fasciculus (ILF), superior longitudinal fasciculus (SLF), corticospinal tract (CST), and uncinate fasciculus (UNC).^21–23^ Thus, we hypothesize that infants with DS would exhibit lower fractional anisotropy (FA) and altered diffusivity in these tracts, reflecting delayed or atypical maturation of neural pathways critical for cognitive and motor function.

By utilizing advanced neuroimaging methods and ensuring a comprehensive assessment, the study aims to make a first attempt to characterize WM microstructure in infants with DS, marking a crucial step in understanding the neurodevelopmental trajectory of DS from its earliest stages.

## Methods

### Study design

This cross-sectional study was reported following STROBE reporting guidelines. Ethical approval was obtained from institutional review boards at all sites that relied on a parent IRB at Washington University in St. Louis, and written informed consent was obtained from each participant’s parent.

### Participants

DS infants were reported by parents to have a diagnosis of trisomy 21 (i.e., not partial or mosaic trisomy 21) and recruited as part of the IBIS-DS study. Control infants without DS, recruited as part of two IBIS infant studies (IBIS-DS and IBIS-Early Prediction), met one of two criteria. Either they had a TD older sibling and no sibling history of autism or neurodevelopmental disorders. Otherwise, if they had an older sibling with an autism diagnosis, they were themselves confirmed not to meet clinical best estimate criteria for an autism diagnosis at 24 months of age, based on DSM-IV-TR and DSM-5-TR. A background on the IBIS infant studies and detailed exclusion criteria are available in **eMethods in Supplement 1**.

### MRI Acquisition and preprocessing

MRI scans were acquired during natural sleep on identical Siemens Prisma 3T scanners with a 32-channel head coil. Diffusion-weighted images (DWIs) were acquired in anterior-posterior (AP) and posterior-anterior (PA) phase-encoding directions, with 102 DWI volumes acquired for each AP and PA: 8 b=0, 20 b= 400, 37 b=1500, 37 b=3000, TR= 3222ms, TE=89.20ms, 1.5mm^3^ voxel, TA=12min19s. Tensors metrics were computed using only b=400 and 1500 shells, while NODDI metrics utilized all available shells. MRI processing and quality control steps are described in **eMethods in Supplement 1**.

A susceptibility artifact in the AP phase affecting the temporal poles was found in 50 scans **(eFigure 1)**. Steps taken to remove the artifact from the analyses are detailed in **eMethods in Supplement 1**.

**Figure 1.**
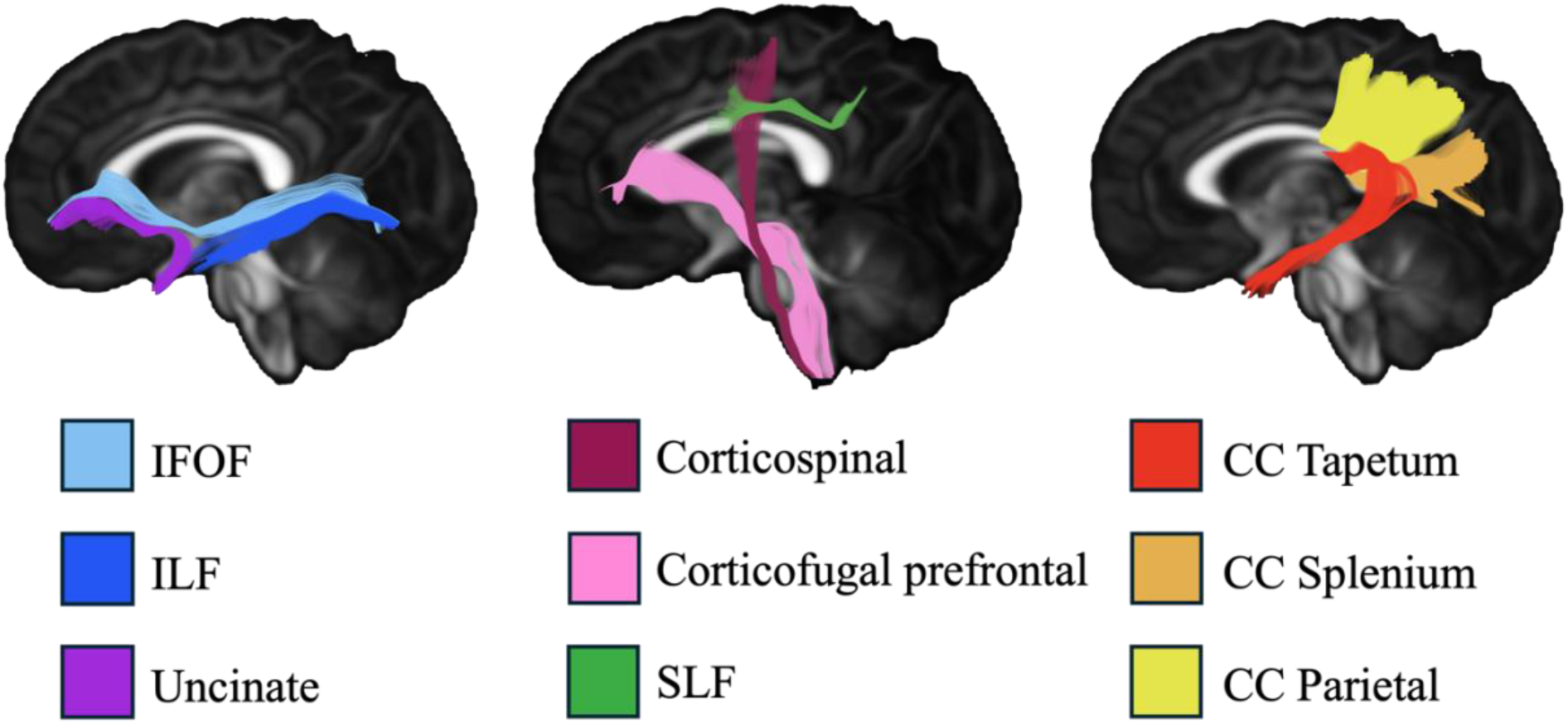
Fiber tractography of the examined tracts. Abbreviations: IFOF, inferior fronto-occipital fasciculus; ILF, inferior longitudinal fasciculus; SLF, superior longitudinal fasciculus; CC, corpus callosum.

### WM Tract Measurements

Metrics of WM microstructure were extracted via an extended UNC-NAMIC automated fiber analysis framework (the detailed process is available in **eMethods in Supplement 1**).^24,25^ Drawing on previous findings in older children^21,23^, we examined 6 intrahemispheric tracts bilaterally—corticofugal prefrontal, CST, IFOF, ILF, SLF II, and UNC—and 3 interhemispheric tracts: the parietal portion, splenium, and tapetum of the corpus callosum (CC). All primary analysis tracts are displayed in **Figure 1**.

**Table 1** outlines DTI and NODDI parameters and their interpretations.

**Table 1.** Definitions of the DTI and NODDI parameters and their interpretation in infants with Down syndrome.

| *Metric* | *Definition of Measurement* | *Interpretation in Infants with Down syndrome* |
| --- | --- | --- |
| **DTI Metrics** |  |  |
| Fractional Anisotropy (FA) | A scalar value between 0 and 1 that quantifies the degree of anisotropy of water diffusion; 0 indicates isotropic diffusion, and 1 indicates fully anisotropic diffusion.<sup>26</sup> | Decreased FA values suggest a delay/disruption in the structural integrity of white matter.<sup>6-8,23</sup> |
| Axial Diffusivity (AD) | The magnitude of water diffusion parallel to the tract.<sup>27</sup> | Decreased AD suggests delayed/disrupted axonal organization and elongation.<sup>28-30</sup> |
| Radial Diffusivity (RD) | The magnitude of water diffusion perpendicular to the tract.<sup>27</sup> | Increased RD suggests delayed/disrupted myelination.<sup>27,31-33</sup> |
| Mean Diffusivity (MD) | The average amount of diffusion occurring within a single voxel.<sup>23</sup> | Increased MD suggests delayed/disrupted tissue organization.<sup>6,34,35</sup> |
| **NODDI Metrics** |  |  |
| Neurite Density Index (NDI) | Reflects the fraction of tissue volume occupied by neurites (axons and dendrites).<sup>36</sup> | Decreased NDI suggests a delayed/disruption in neurite density.<sup>37,38</sup> |
| Orientation Dispersion Index (ODI) | The variability in neurite orientation, ranging from 0 (perfectly aligned) to 1 (randomly in all directions).<sup>36</sup> | Increased ODI suggests greater dispersion and fanning of neurites.<sup>39-41</sup> |
| Free Water Fraction (FWF) | The fraction of diffusion signals explained by isotopically unrestricted water, estimated using a bi-tensor model.<sup>42</sup> | Increased FWF suggests neuroinflammation.<sup>43,44</sup> |

### Statistical Analyses

All statistical analyses were performed using general linear models (GLMs) in JMP 17 Pro.

#### Primary Analyses

For each tract, a multivariate analysis of variance (MANOVA) was conducted comparing the average values of FA, RD, AD, NDI, and ODI between the DS and control groups. MD was excluded to avoid multicollinearity with AD and RD in MANOVA, while FWF was excluded due to its sensitivity to partial volume effects, particularly in tracts close to the cerebrospinal fluid (CSF), like the CC.

For tracts showing significant group differences in MANOVA, follow-up univariate analyses of variance (ANOVA) were conducted to identify the specific diffusion parameters significantly different between the two groups. All analyses were covaried by age-at-assessment in days, sex, and scan-motion quantification (defined as the number of diffusion volumes with significant artifacts or head motion greater than 2mm). To correct for multiple comparisons, Bonferroni correction was applied across all tests, accounting for 15 comparisons in MANOVA (*corrected p<0.003*) and 5 diffusion parameter comparisons in ANOVA (*corrected p<0.01*). All reported *p-values* from ANOVA tests in the Results section are corrected.

#### Secondary Analyses

For tracts identified as significant in ANOVA, along-tract analyses were conducted using the Functional Analysis of Diffusion Tensor Tract Statistics (FADTTS) toolbox^45^ and its corresponding graphical user interface, FADTTSter.^46^ Along-tract analyses were performed for each diffusion parameter (FA, RD, AD, NDI, ODI) identified as significant in prior ANOVA tests, providing a finer-grained understanding of where differences occurred in the tract. Results were assessed visually and statistically to interpret region-specific differences in diffusion properties.

Finally, exploratory full-brain analyses were performed to examine all diffusion parameters (FA, AD, RD, MD, NDI, ODI, and FWF) across all 51 tracts (listed in **eTable 1**), and were conducted without correcting for multiple comparisons, providing a broader view of potential differences between groups.

## Results

### Demographics

A total of 49 DS and 37 control infants were included. No significant differences were observed in sex (*X*^2^=0.61, *p*=0.43), age-at-assessment in days (*p*=0.12), and scan motion quantification (*p*=0.6). Gestational age was slightly higher in DS group (267 ± 9.5 days) than in control group (272 ± 9 days; *p*=0.015). Maternal age at birth was also significantly higher in DS group (36.84 ± 4.54 years) than in control group (34.16 ± 3.02 years; *p*=0.004), consistent with the increased incidence of DS with maternal age. A full summary of the demographics can be found in **Table 2**.

**Table 2.** Participant demographics by group. (\*\*\**p*-value is <0.0001, \*\**p*-value is 0.01 - 0.001, \**p*-value is 0.05 - 0.01).

|  | *Down syndrome (DS)* | *Controls* | *Group Comparison* |
| --- | --- | --- | --- |
| **N** | 49 | 37 |  |
| **Sex, n (%)** | Female: 28 (57.14)<br>Male: 21 (42.86) | Female: 18 (48.65)<br>Male: 19 (51.35) | $\chi^2=0.611$, $p$-value=0.4343 |
| **Age-at-assessment in days, Mean (SD)** | 207.31 (25.10) | 199.62 (19.09) | t-test, $p$-value=0.1242 |
| **Gestational Age in days, Mean (SD)** | n=39, 266.63 (9.46) | n=33, 272.09 (9.05) | t-test, $p$-value=0.0152* |
| **Scan-motion quantification, Mean (SD)** | 10.20 (10.34) | 8.92 (12.27) | t-test, $p$-value=0.6 |
| **Maternal Age at birth in years, Mean (SD)** | n=38, 36.84 (4.54) | n=36, 34.16 (3.02) | t-test, $p$-value=0.004** |
| **Paternal Age at birth in years, (Mean) (SD)** | n=37, 37.71 (6.44) | n=36, 36.03 (4.96) | t-test, $p$-value=0.2171 |
| **Maternal Education, Mean (SD)** | 4.24 (1.30) | 4.6 (1.16) | t-test, $p$-value=0.2156 |
| 1. Some high school | 0 (0) | 0 (0) |  |
| 2. High school graduate | 3 (6.12) | 0 (0) |  |
| 3. Some college | 8 (16.33) | 6 (16.22) |  |
| 4. College graduate | 15 (30.61) | 15 (40.54) |  |
| 5. Some grad school | 1 (2.04) | 1 (2.7) |  |
| 6. Graduate degree | 11 (22.45) | 13 (35.14) |  |
| NA. Not available | 11 (22.45) | 2 (5.41) |  |
| **Paternal Education, Mean (SD)** | 4.027 (1.4237) | 4.4722 (1.5021) | t-test, $p$-value=0.1978 |
| 1. Some high school | 0 (0) | 0 (0) |  |
| 2. High school graduate | 7 (14.29) | 4 (10.81) |  |
| 3. Some college | 5 (10.2) | 7 (18.92) |  |
| 4. College graduate | 15 (30.61) | 9 (24.32) |  |
| 5. Some grad school | 0 (0) | 0 (0) |  |
| 6. Graduate degree | 10 (20.41) | 16 (42.24) |  |
| NA. Not available | 12 (24.49) | 1 (2.7) |  |
| **Household Income, Mean (SD)** | 5.8 (1.86) | 6.08 (1.36) | t-test, $p$-value=0.4653 |
| 1. less than 25K | 2 (4.08) | 0 (0) |  |
| 2. 25K-35K | 0 (0) | 0 (0) |  |
| 3. 35K-50K | 1 (2.04) | 1 (2.70) |  |
| 4. 50K-75K | 6 (12.25) | 4 (10.81) |  |
| 5. 75K-100K | 2 (4.08) | 6 (16.22) |  |
| 6. 100K-150K | 11 (22.45) | 12 (32.43) |  |
| 7. 150K-200K | 6 (12.25) | 6 (16.22) |  |
| 8. over-200K | 7 (14.29) | 7 (18.92) |  |
| NA. Not available | 14 (28.57) | 1 (2.70) |  |

### Analysis of Variance of Diffusion Parameter Averages

Significant group differences were found between DS and control groups on MANOVA in the tapetum and parietal portions of the CC, as well as bilateral CST, IFOF, ILF, SLF II, and UNC. Left SLF II (*F*=12.16, *p*<.0001), right SLF II (*F*=10.73, *p*<.0001), left IFOF (*F*=10.57, *p*<.0001) and parietal CC (*F*=8.51, *p*<.0001) showed the strongest statistical differences on MANOVA. The splenium of the CC and bilateral corticofugal prefrontal tracts showed no group differences. **eTable 2** details the MANOVA results.

On ANOVA, several association tracts in DS infants showed patterns consistent with reduced structural integrity and neurite density, as evidenced by reduced FA and NDI, and delayed maturation indicated by increased RD. IFOF showed bilateral reductions in FA (Left: *β*=0.012, Cohen’s-*d*=-1.37, *p*<.0001; Right: *β*=0.0087, Cohen’s-*d*=-0.95, *p*<.0001), and NDI (Left: *β*=0.0069, Cohen’s-*d*=-0.69, *p*<.0001; Right: *β*=0.0077, Cohen’s-*d*=-0.69, *p*<.0001), and increase in RD (Left: *β*=-0.000014, Cohen’s-*d*=0.94, *p*<.0001; Right: *β*=-0.000013, Cohen’s-*d*=0.79, *p*<.0001) in DS group. SLF II showed bilateral reductions in FA (Left: *β*=0.012, Cohen’s-*d*=-1.23, *p*<.0001; Right: *β*=0.0082, Cohen’s-*d*=-0.811, *p*=0.004), and NDI (Left: *β*=0.0093, Cohen’s-*d*=-0.85, *p*<.0001; Right: *β*=0.0088, Cohen’s-*d*=-0.8, *p*<.0001), and increase in RD bilaterally (Left: *β*=-0.000018, Cohen’s-*d*=1.04, *p*<.0001; Right: *β*=-0.000012, Cohen’s-*d*=0.66, *p*<.0001) in DS group.

IFOF and UNC also showed increased neurite dispersion and fanning bilaterally reflected by elevated ODI in DS group (Left IFOF: *β*=-0.0058, Cohen’s-*d*=1.21, *p*<.0001; Right IFOF: *β*=-0.0039, Cohen’s-*d*=0.79, *p*=0.004; Left UNC: *β*=-0.0038, Cohen’s-*d*=0.74, *p*=0.023; Right UNC: *β*=-0.005, Cohen’s-*d*=0.94, *p*=0.001).

The parietal portion of the CC and bilateral CST demonstrated patterns of increased axonal integrity as indicated by elevated AD (Parietal CC: *β*=-0.000013, Cohen’s-*d*=0.63, *p*=0.009; Left CST: *β*=−0.000019, Cohen’s-*d*=0.84, *p*<.0001; Right CST: *β*=−0.000014, Cohen’s-*d*=0.54, *p*=0.035). The tapetum of the CC showed no significant differences after correcting for multiple comparisons.

Figure 2 presents violin and box plots of the significant diffusion metrics in the bilateral CST and IFOF. **Table 3** details the ANOVA results for tracts found significant on MANOVA.

**Figure 2.**
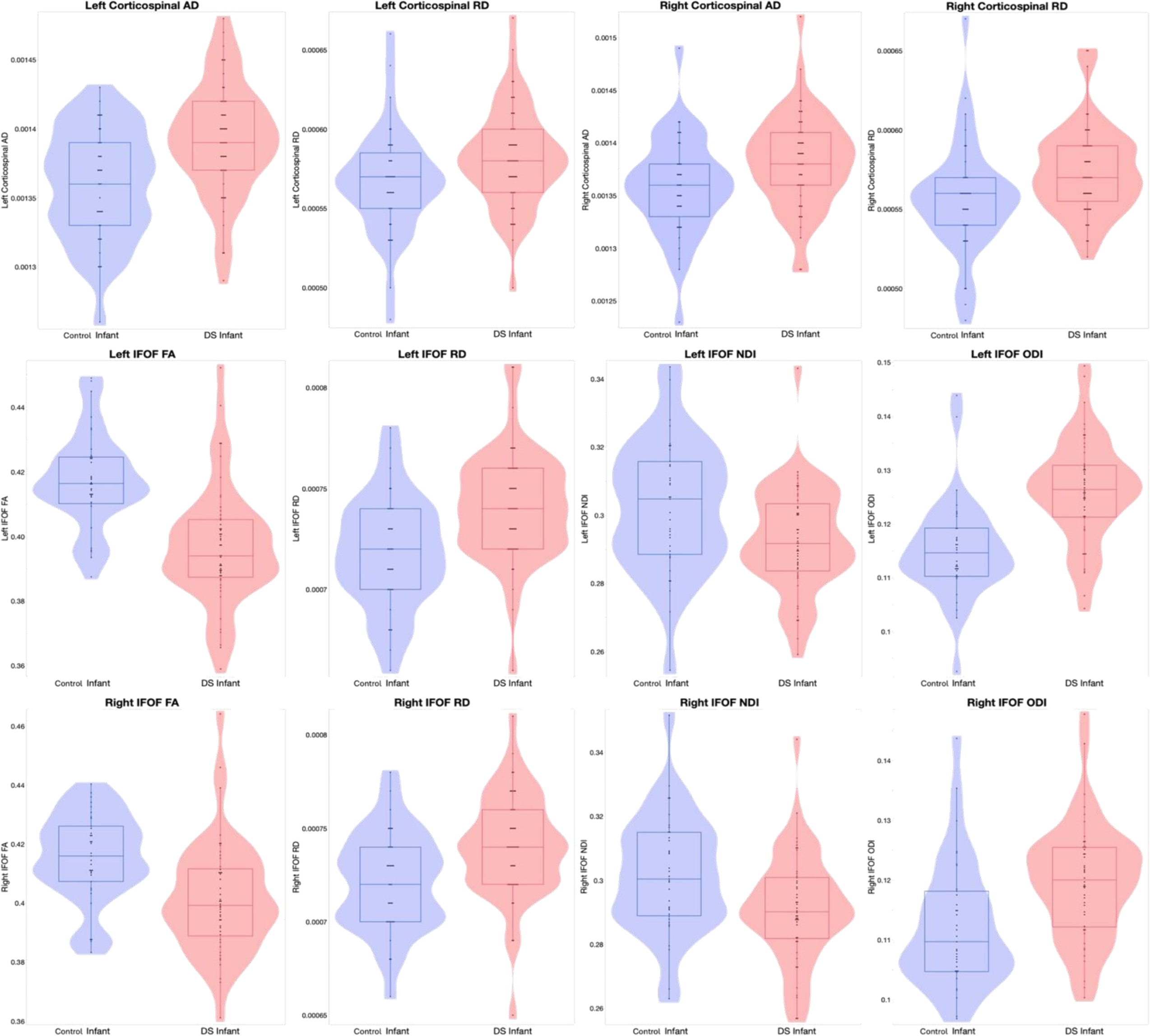
Violin and box plots of the significant diffusion metrics in the bilateral CST and IFOF. Abbreviations: IFOF, inferior fronto-occipital fasciculus; CST, corticospinal tract. FA, fractional anisotropy; AD, axial diffusivity; RD, radial diffusivity; NDI, neurite density index; ODI, orientation dispersion index.

**Table 3.**
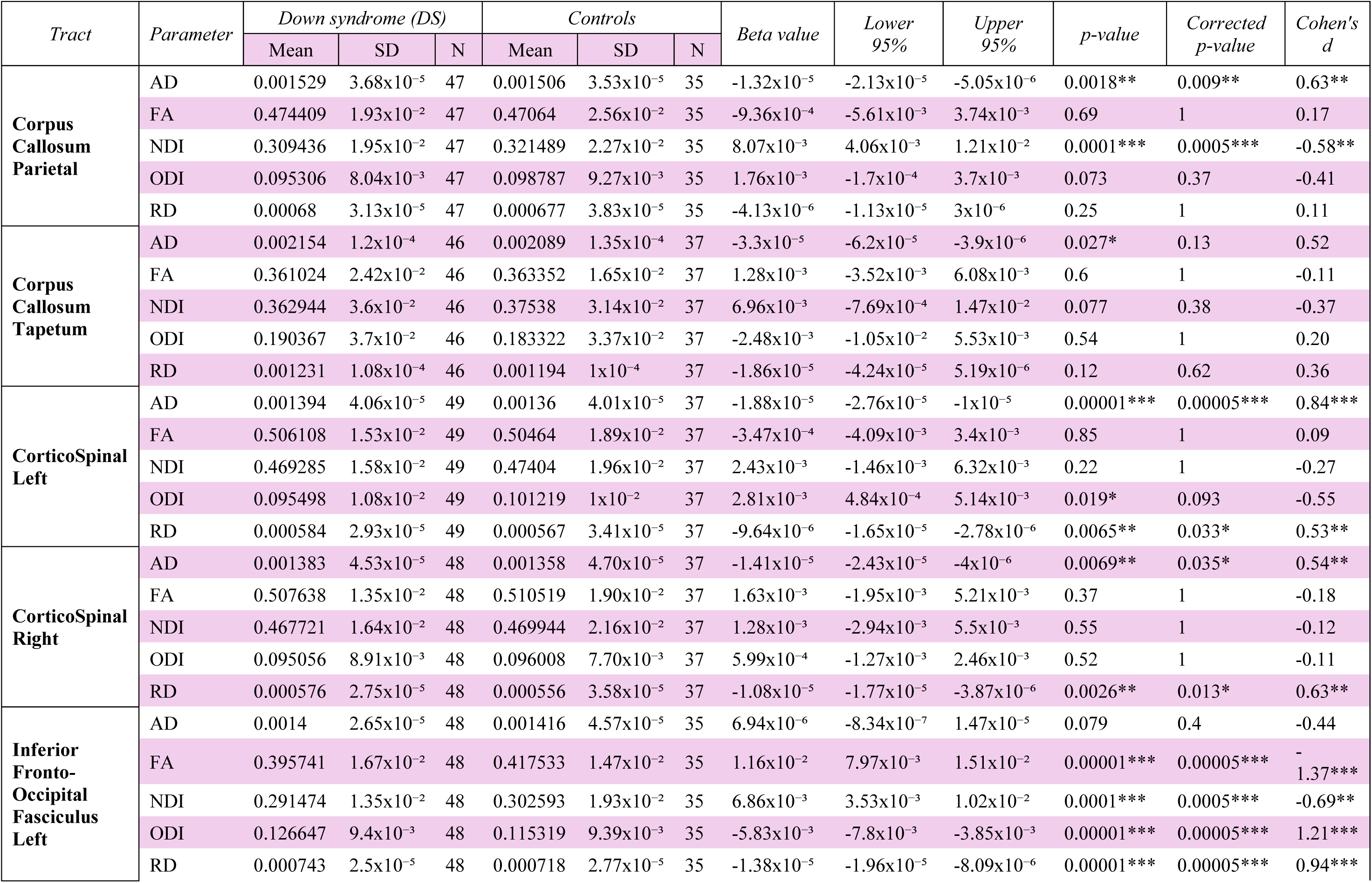
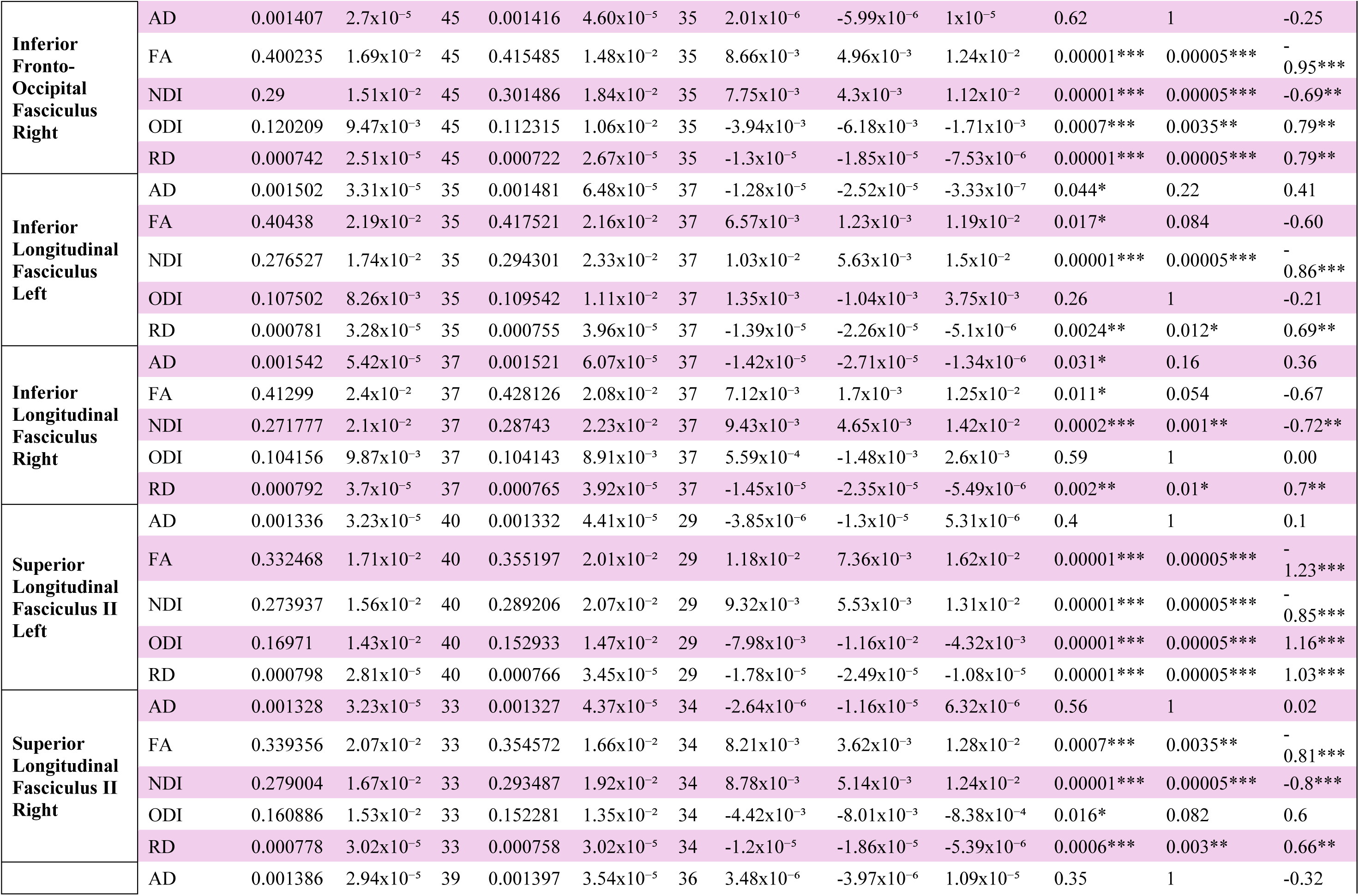
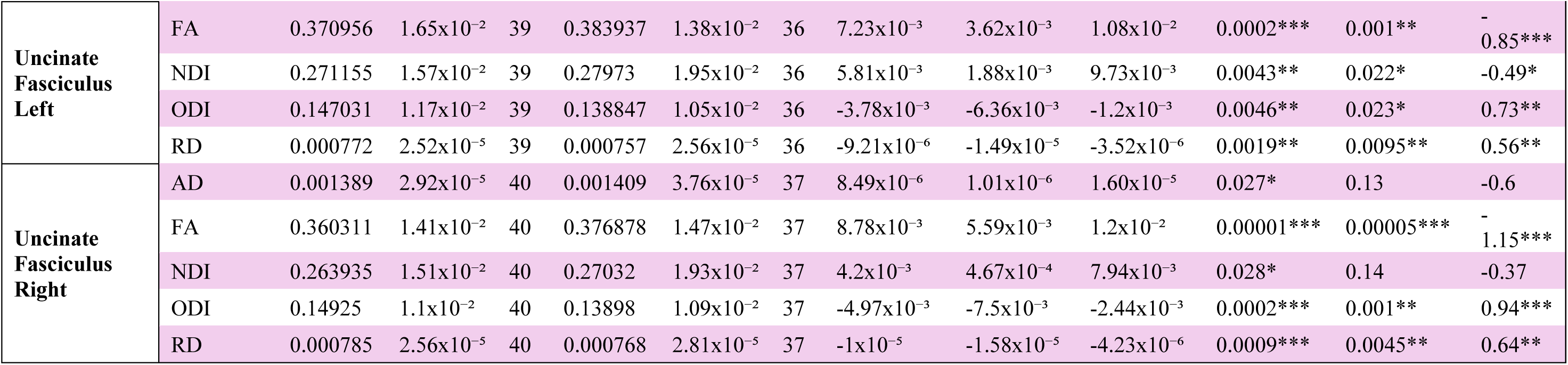
Results of univariate analyses of variance comparing DTI and NODDI parameters in the significant tracts between DS and control infants. (\*\*\**p*-value is <0.0001, \*\**p*-value is 0.01 - 0.001, \**p*-value is 0.05 - 0.01; Cohen’s d: *small effect is 0.2 – 0.5, **medium effect is 0.5 – 0.8, ***large effect is ≥ 0.8).

### Along-Tract Analyses

We conducted follow-up along-tract analyses for the diffusion parameters found significant in ANOVA to identify spatially-specific differences along the trajectories of fiber tracts.

Along-tract analysis of the bilateral CST revealed significant group differences in RD in the region between the midbrain and the internal capsule. The left ILF showed significant differences in RD in the ventral portion of the tract within the temporal lobe, while the left SLF II demonstrated differences in NDI and RD in the ventral portion of the tract as it extends into the frontal lobe. The right IFOF revealed consistent differences in FA, RD, and NDI across the frontal, temporal and parietal portions of the tract. Lastly, the right UNC exhibited differences in FA and ODI in the frontal portion of the tract. **e**Figure 2 visualizes the significant *p*-values along the tracts.

### Exploratory Analyses

Exploratory analyses revealed distinct patterns in diffusion parameters between DS and control infants. FA values were lower in DS infants across all significant tracts except for the bilateral optic tracts, which exhibited higher FA values. MD values were consistently higher in DS infants, while both AD and RD values were higher in DS infants, with the exception of the left cingulate gyrus of the cingulum (CGC) for AD and the right optic tract for RD.

NDI values were consistently lower in DS infants across all significant tracts. ODI values varied across tracts, with some showing increases and others decreases in DS infants. FWF values were lower in DS infants in all significant tracts except for the CST bilaterally and the right hippocampal part of the cingulum (HCG).

Notably, the bilateral optic tracts were the only tracts to demonstrate higher FA and AD values, alongside lower RD and ODI values, suggesting preserved or distinct WM microstructure.

Full statistical results across all tracts are presented in **eTable 3**.

## Discussion

This is the first study to examine the WM microstructure in infants with DS. Findings reveal consistent patterns of delayed WM maturation, characterized by regional differences in neurite density, axonal growth, and myelination. DS infants exhibited reduced microstructural coherence and delayed myelination across multiple intrahemispheric tracts, including the bilateral IFOF, SLF, and UNC, indicated by significantly lower FA and elevated RD compared to their control counterparts. Additionally, DS infants exhibited significantly elevated RD in motor pathways such as the bilateral CST. These findings align with prior DS research in early childhood to adulthood.^6,21–23^

Exploratory analyses showed increased MD in specific tracts, including the frontoparietal portion of the left arcuate fasciculus, left SLF II, and left corticothalamic motor and premotor pathways. In typical development, myelination progresses posterior-to-anterior and caudal-to-rostral, with occipital/parietal lobes myelination occurring between 4– 6 months and frontal/temporal lobes between 6–8 months.^47–50^ As such, it is expected that tracts located in the frontal regions would show relatively high MD during this developmental stage. The observation that DS infants exhibited higher MD in these tracts, compared to controls of the same age, suggests that the typical timeline of myelination is delayed or disrupted in DS infants, reflecting slower and less complete myelination.

Significant elevation in AD in DS infants was observed in both inter- and intrahemispheric tracts, including the CST, ILF, the parietal and tapetum portions of the CC, indicating alterations in axonal development during early growth. Romano et al.^22^ reported elevated AD in the CST of young adults with DS, as well as forceps major and minor. In contrast, the lower AD and FA and higher RD in the left CGC and right UNC indicate regional delays in axonal development and myelination, highlighting differential vulnerability of pathways associated with attention, memory and emotional regulation.^6,51–53^

NDI was significantly reduced in DS infants across most major WM tracts, including the SLF II, IFOF, ILF, and the majority of the CC except the tapetum. Previous studies indicate that NDI typically reflects neurite density, which generally increases during the first two decades of life.^40,54^ Lower NDI in DS infants suggests a slower rate of neurite packing, particularly in axons and dendrites, during this critical developmental window. Consistent with Timmers et al.^55^ findings, NDI demonstrated greater sensitivity, identifying more tracts with lower values compared to FA. Combined with reductions in FA, the lower NDI provides converging evidence of global delays in WM maturation in DS infants.

Changes in ODI further emphasize regional variability in neurite structure. In DS infants, increased ODI in the bilateral IFOF, UNC, and SLF II suggests greater neurite dispersion, aligning with findings by Garic et al.^21^ in school-aged children. This may reflect compensatory reorganization, such as axonal sprouting or delayed pruning, in response to reduced neurite density, as indicated by lower NDI.^56,57^

In our exploratory analyses, significant reductions in FWF were observed in the CC (body, motor, parietal) and corticothalamic motor and right premotor tracts, which also showed concurrent reductions in NDI. This pattern suggests reduced neurite density without evidence of neuroinflammation. While direct evidence for elevated FWF in older individuals with DS is insufficient, increased FWF has been documented in older adults with AlzD.^58,59^ DS is associated with early onset and increased prevalence of AlzD, purportedly due to the triplicated amyloid precursor protein on chromosome 21.^5,60,61^ Thus, increased FWF in older DS individuals may reflect neurodegeneration or inflammation similar to that seen in AlzD. However, the reduced FWF observed in DS infants suggests an absence of inflammatory or degenerative processes at this stage, highlighting the need for longitudinal studies to distinguish the pathological changes in WM across the lifespan.

The optic tracts exhibit a distinct developmental profile, suggesting an early maturation of sensory pathways relative to higher-order systems. Infants with DS exhibited higher FA in the optic tracts bilaterally, consistent with findings from Gunbey et al.^23^, who observed elevated FA in the right optic tract of two-year-old children with DS. This increase in FA, alongside elevated AD, suggests a more linear and organized axonal structure, indicative of preserved neurite density in this sensory pathway. Furthermore, reduced ODI in the optic tracts aligns with these findings, reflecting less dispersion and greater coherence in fiber orientation, consistent with early maturation typical of the visual pathway.^62,63^ However, this early structural advantage does not necessarily translate to functional gains over time, as studies show that visual acuity plateaus after two years in children with DS,^64^ while it continues to improve in TD peers.^65,66^

Along-tract analyses offer a detailed spatially specific assessment of WM changes compared to whole-tract average diffusion parameters.^67,68^ Findings revealed localized alterations in the bilateral CST, left SLF II and ILF, and right IFOF and UNC. However, the absence of significant along-tract findings in other tracts, despite group differences detected in average parameter analyses, could be attributed to the increased number of comparisons required by along-tract approaches, which demand a larger sample size to detect significant localized changes along the tracts. Our findings demonstrate that WM microstructural alterations emerge early in DS infants, mirroring patterns observed in later developmental stages,^6–8,21–23,60^ underscoring the importance of exploring this underrepresented field in DS neurodevelopment. Early interventions and therapies have been shown to improve outcomes in individuals with DS,^69^ and WM may serve as a valuable biomarker for monitoring these interventions, as it has shown to change in response to treatment in children and adults.^70^ Notably, a prior clinical trial reported improvements in WM connectivity following treatment in children with autism.^71^ This study represents a crucial foundation for understanding the neurobiology of DS and identifying early WM alterations that may inform optimal windows for mechanistically-derived interventions aimed at improving long-term outcomes.

A key strength of our study is its larger sample of DS and control infants compared to previous studies in toddlers, the largest of which^23^ included only 10 DS and 8 control individuals. The larger sample size enhances the reliability and generalizability of our findings. The use of multishell diffusion imaging allowed us to obtain and examine both DTI and NODDI data in DS infants. NODDI distinguishes intra-neurite, extra-neurite, and CSF compartments, estimating neurite density and orientation dispersion.^40,72,73^ These measures complement traditional DTI, helping disentangle the effects of axonal density, myelination, and neurite organization.^74,75^

### Limitations and future directions

This study has a few limitations. A larger sample size would improve the ability to detect smaller effects in along-tract analyses. Additionally, the observed susceptibility artifact, as described in the **eMethods in Supplement 1**, necessitated a more stringent definition of certain tracts, which could have influenced the findings in the affected tracts. Future research should include longitudinal follow-up to chart WM microstructure development in DS from infancy to childhood, as well as examining how WM microstructure in infancy correlates with behavioral, language, and cognitive outcomes during childhood, potentially providing a predictive tool for these developmental outcomes.

## Conclusion

Our findings reveal distinct patterns of delayed WM maturation in DS infants, marked by reduced myelination, lower neurite density, and increased neural dispersion in fibers critical for higher-order cognitive and motor functions, providing an early window into the microstructural abnormalities that may underlie later cognitive and motor delays.

By employing advanced diffusion imaging techniques, this study examines WM microstructure in DS infants, addressing a significant gap in research that has largely focused on older children and adults.

These findings lay a foundation for future longitudinal studies to explore how early WM alterations relate to cognitive, behavioral, and motor outcomes in DS, and will be essential for identifying critical windows for targeted clinical interventions aimed at supporting WM maturation and mitigating developmental delays.

## Supporting information

Supplemental eTable 3

## Data Availability

All data produced in the present study are available upon reasonable request to the authors.

## Acknowledgements

**Information on author access to data:** Dr. Azrak had full access to all of the data in the study and takes responsibility for the integrity of the data and the accuracy of the data analysis.

**Disclosure of potential conflicts of interest:** Dr. Robert McKinstry holds stock options in Turing Medical and serves as a paid member of its medical advisory board. He also receives travel, lodging, and meal support from Siemens Healthcare and Radiaction, as well as meal support from Hyperfine.

**Funding/Support:** This study was supported by grants from the National Institutes of Health (R01HD088125, K01HD109445, K23HD112809, R01HD055741, R01MH118362, R01MH118362–02S1, P50HD103573-8084, T32HD040127, K01MH122779).

**Role of the Funder/Sponsor:** The funder had no role in the design and conduct of the study; collection, management, analysis, and interpretation of the data; preparation, review, or approval of the manuscript; and decision to submit the manuscript for publication.

**Previous presentations:** Preliminary results were presented at the Flux 2024 Congress in Baltimore, MD on September 29, 2024. The work is not being considered for any future meetings.

**Use of artificial intelligence:** The authors used ChatGPT (version 4o, accessed via ChatGPT Plus) by OpenAI for writing assistance in the development of the manuscript. This tool was used between November 2024 and February 2025 to refine wording, enhance clarity, and improve the readability of various sections. At no point did the AI generate original scientific content, conduct data analysis, or make independent claims about the study findings. The authors take full responsibility for the accuracy, integrity, and originality of the manuscript and its content.

## Notes

### Author Declarations

IRBs of the Childrens Hospital of Philadelphia, The University of Minnesota, The University of North Carolina at Chapel Hill, The University of Washington relying on a parent IRB at Washington University in St. Louis all gave ethical approval for this work.

### Summary of Updates

The author list has been updated to include an essential contributor, and the license has been changed to CC-BY. No other changes have been made to the manuscript or its supplemental files.

