## Supplemental Material for "Early White Matter Microstructure Alterations in Infants with Down Syndrome"

**eMethods.** Supplementary Methods

**eFigure 1. An example of the susceptibility artifact is seen bilaterally in the temporal poles on high b-value (A) and low b-value (B) DWI.**

**eTable 1. Index of all tracts examined in secondary analyses.**

**eTable 2. Results of Multivariate analyses of variance comparing DTI and NODDI parameters in the tracts of interest between DS and control infants**. ($^{***}p$-value is <0.0001, $^{**}p$-value is 0.01 - 0.001, $^{*}p$-value is 0.05 - 0.01).

**eFigure 2. Visualization of the significant *p*-values along the tracts generated using Slicer.** (A) displays significant regions of RD in the bilateral CST, (B) highlights significant RD in the left SLF II and ILF, and (C) shows significant FA in the right IFOF and UNC. Abbreviations: IFOF, inferior fronto-occipital fasciculus; ILF, inferior longitudinal fasciculus; SLF, superior longitudinal fasciculus; CST, corticospinal tract; UNC, uncinate fasciculus. FA, fractional anisotropy; RD, radial diffusivity.

**eTable 3.** Full statistical results across all tracts**.**

### eMethods

**IBIS Infant Studies Background:**

The study is part of the Infant Brain Imaging Study (IBIS), an ongoing multisite longitudinal study, collecting developmental behavioral and neuroimaging data on infants with neurodevelopmental conditions. The IBIS–DS studies infants with and without DS between the ages of 6 and 24 months. The IBIS–Early Prediction (IBIS-EP) studies infants at elevated familial likelihood for ASD between the ages of 6 and 24 months.

Data collection occurred at five sites: The Children's Hospital of Philadelphia (CHOP), The University of Minnesota (UMN), The University of North Carolina at Chapel Hill (UNC), The University of Washington (UW), and Washington University in St. Louis (WashU). Infants were recruited, scanned, and examined between March 2019 and May 2024.

**Exclusion Criteria:**

Exclusion criteria for all infants included: a diagnosis or physical signs of known genetic conditions (other than DS in DS infants), significant medical conditions the could impact growth, development, cognition, or sensory function (except for conditions commonly associated with DS in DS infants), birth weight <2500 grams, gestational age <34 weeks for DS infants or <36 weeks for control infants, history of significant perinatal complications, prenatal exposure to neurotoxins, maternal gestational diabetes requiring medication, contraindication for MRI, and families whose primary language is not English, due to requirements for cognitive assessments.

**MRI Processing and Quality Control:**

All native data underwent automated quality control (QC) using DTIPrep1.2.9[1] to remove any poor-quality DWI volumes. A preliminary visual QC using 3D Slicer 5.1.0 was then performed by a trained rater who visually assessed the quality of the remaining volumes, excluding any scan with significant motion artifact, consistent signal dropout, or surviving volumes below a predetermined threshold (80%). For participants with multiple series, the rater selected the highest-quality series.

Following automatic and preliminary QC, the remaining volumes were corrected for susceptibility, motion, and eddy current artifacts jointly via topup and eddy_openmp in FSL 6.0.3.[2] In the same procedure, the remaining artifact/outlier data were corrected/interpolated in FSL 6.0.3.[3] Brain masks were automatically generated as part of this procedure as these are necessary for the FSL processing. The automatic brain masks were computed via FSL bet applied to the (susceptibility corrected) average baseline/B0 image. All brain masks were reviewed and edited by trained raters.

A final visual QC was conducted by a trained rater who assessed the individual DWI volumes using DMRIPrep 0.5.0. DWI volumes with significant motion artifacts or signal dropouts were excluded. The rater also evaluated each scan's tractography based on the general expected anatomy, tract symmetry, and major tract integrity. Only scans with expected tractography and >80% surviving gradients passed.

**DTI Processing:**

A study-specific diffeomorphic, unbiased DTI atlas was created using the UNC-Utah National Alliance for Medical Image Computing DTI framework.[4] Brain-masked Diffusion Tensor Images (weighted least squares estimation of tensors) of 70 subjects were used to create the atlas. Then, each individual subject DTI were mapped to atlas space using nonlinear, diffeomorphic pair-wise registration, with visual inspection to verify registration accuracy.

**Susceptibility Artifact Removal:**

To remove the artifact from the analyses, the region containing the artifact was manually segmented in 10 individual DWI scans, with the mask then propagated to the DWI atlas for the entire sample. Any segments of tracts affected by the artifact were excluded from the analyses. Tracts affected were the tapetum of the corpus callosum, bilateral ILFs, bilateral fornices, and bilateral UNCs. There was no significant group difference in the occurrence rate of the artifact ($X^2$=1.44, $p$=0.23).

**White Matter Metrics Extraction:**

Fiber tractography was performed using AutoTract followed by manual refinement based on established fiber definitions.[5] The resulting deformation fields mapped atlas fibers to individual subject spaces, where diffusion tensor and NODDI metrics were extracted at equidistant points along each fiber tract. Diffusion tensors were estimated via a weighted least squares approach in Dmriprep[6], DTI properties—FA, AD, RD, MD—were extracted from the estimated tensors. NODDI properties—neurite density index (NDI) and orientation dispersion index (ODI)—were generated using Accelerated Microstructure Imaging via Convex Optimization (AMICO).[7]

For quality control, we excluded subjects whose tract-specific FA profiles correlated poorly ($r < 0.70$) with the population average, as this typically indicates suboptimal DTI-to-atlas alignment. Final tract metrics were computed by averaging the respective diffusion parameters along each fiber tract.

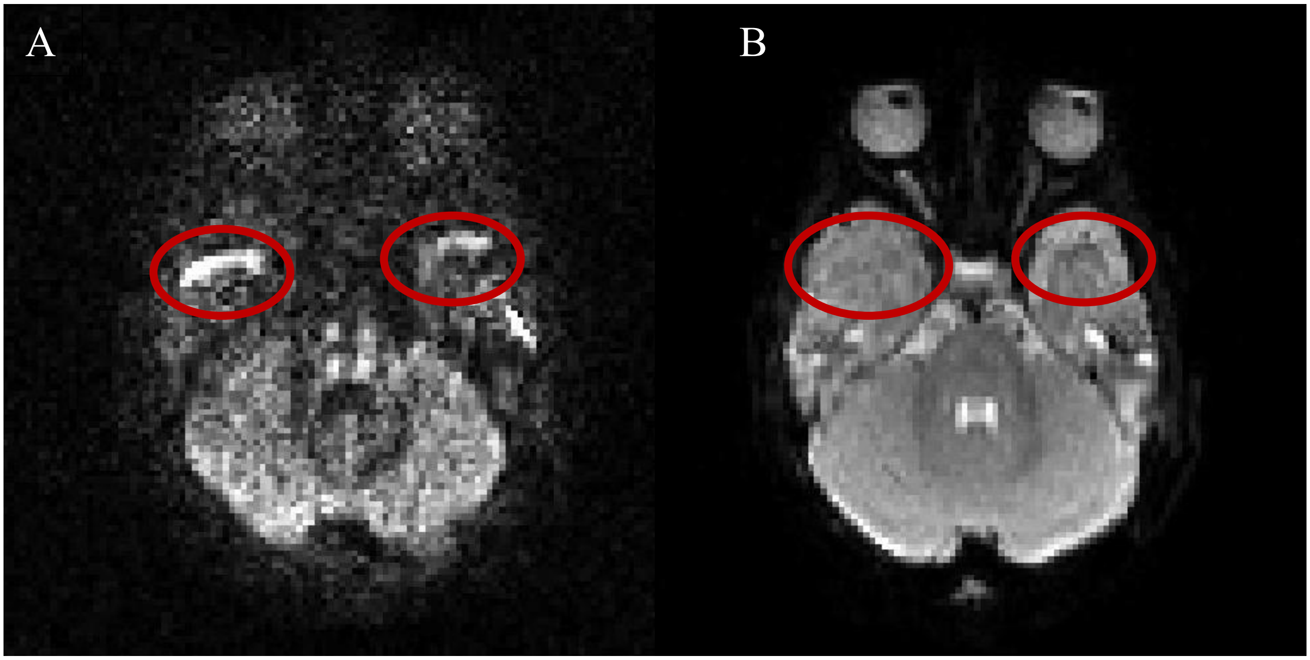

**eFigure 1. An example of the susceptibility artifact is seen bilaterally in the temporal poles on high b-value (A) and low b-value (B) DWI.**

| *Tracts* | |
|---|---|
| Arcuate Fasciculus FrontoParietal Left | Corticothalamic Motor Left |
| Arcuate Fasciculus FrontoParietal Right | Corticothalamic Motor Right |
| Arcuate Fasciculus FrontoTemporal Left | Corticothalamic Parietal Left |
| Arcuate Fasciculus FrontoTemporal Right | Corticothalamic Parietal Right |
| Arcuate Fasciculus TemporoParietal Left | Corticothalamic PreFrontal Left |
| Arcuate Fasciculus TemporoParietal Right | Corticothalamic PreFrontal Right |
| Cingulate Gyrus part of the Cingulum Left | Corticothalamic PreMotor Left |
| Cingulate Gyrus part of the Cingulum Right | Corticothalamic PreMotor Right |
| Corpus Callosum Body | Corticothalamic Superior Left |
| Corpus Callosum Genu | Corticothalamic Superior Right |
| Corpus Callosum Motor | Fornix Left |
| Corpus Callosum Parietal | Fornix Right |
| Corpus Callosum PreMotor | Hippocampal part of the Cingulum Left |
| Corpus Callosum Splenium | Hippocampal part of the Cingulum Right |
| Corpus Callosum Tapetum | Inferior Fronto-Occipital Fasciculus Left |
| Corticofugal Motor Left | Inferior Fronto-Occipital Fasciculus Right |
| Corticofugal Motor Right | Inferior Longitudinal Fasciculus Left |
| Corticofugal Parietal Left | Inferior Longitudinal Fasciculus Right |
| Corticofugal Parietal Right | Optic Radiation Left |
| Corticofugal PreFrontal Left | Optic Radiation Right |
| Corticofugal PreFrontal Right | Optic Tract Left |
| Corticofugal PreMotor Left | Optic Tract Right |
| Corticofugal PreMotor Right | Superior Longitudinal Fasciculus II Left |
| CorticoSpinal Left | Superior Longitudinal Fasciculus II Right |
| CorticoSpinal Right | Uncinate Fasciculus Left |
| | Uncinate Fasciculus Right |

**eTable 1. Index of all tracts examined in secondary analyses.**

**eTable 2. Results of Multivariate analyses of variance comparing DTI and NODDI parameters in the tracts of interest between DS and control infants.** ($^{***}$*p*-value is <0.0001, $^{**}$*p*-value is 0.01 - 0.001, $^{*}$*p*-value is 0.05 - 0.01).

| *Tract* | *Test* | *Multivariate Analysis of Variance* | | | | *Cohort* | |
|---|---|---|---|---|---|---|---|
| | | Value | Approx. F | *p*-value | Corrected *p*-value | Exact F | *p*-value |
| **Corpus Callosum Parietal** | Wilks' Lambda | 0.3517668 | 4.5001 | 0.00001*** | 0.00015*** | 8.5062 | 0.00001*** |
| | Pillai's Trace | 0.7936431 | 3.7623 | 0.00001*** | 0.00015*** | | |
| **Corpus Callosum Splenium** | Wilks' Lambda | 0.5177167 | 2.7412 | 0.0001*** | 0.0015** | 1.9091 | 0.1028 |
| | Pillai's Trace | 0.5654121 | 2.5681 | 0.0003*** | 0.0045** | | |
| **Corpus Callosum Tapetum** | Wilks' Lambda | 0.5625489 | 2.3333 | 0.0014** | 0.021* | 5.7214 | 0.0002*** |
| | Pillai's Trace | 0.4940561 | 2.1702 | 0.003** | 0.045* | | |
| **Corticofugal PreFrontal Left** | Wilks' Lambda | 0.5676535 | 2.386 | 0.001** | 0.015* | 1.0374 | 0.4019 |
| | Pillai's Trace | 0.4985278 | 2.278 | 0.0016** | 0.024* | | |
| **Corticofugal PreFrontal Right** | Wilks' Lambda | 0.574091 | 2.3344 | 0.0013** | 0.0195* | 1.369 | 0.2453 |
| | Pillai's Trace | 0.4946746 | 2.2579 | 0.0018** | 0.027* | | |
| **Corticospinal Left** | Wilks' Lambda | 0.5797876 | 2.2894 | 0.0017** | 0.0255* | 6.0233 | 0.00001*** |
| | Pillai's Trace | 0.476954 | 2.1661 | 0.003** | 0.045* | | |
| **Corticospinal Right** | Wilks' Lambda | 0.5869572 | 2.2046 | 0.0027** | 0.0405* | 5.344 | 0.0003*** |
| | Pillai's Trace | 0.4718462 | 2.1131 | 0.004** | 0.06 | | |
| **Inferior Fronto-Occipital Fasciculus Left** | Wilks' Lambda | 0.318121 | 5.081 | 0.00001*** | 0.00015*** | 10.5716 | 0.00001*** |
| | Pillai's Trace | 0.921279 | 4.6083 | 0.00001*** | 0.00015*** | | |
| **Inferior Fronto-Occipital Fasciculus Right** | Wilks' Lambda | 0.4373586 | 3.3478 | 0.00001*** | 0.00015*** | 6.7666 | 0.00001*** |
| | Pillai's Trace | 0.6948693 | 3.1115 | 0.00001*** | 0.00015*** | | |
| **Inferior Longitudinal Fasciculus Left** | Wilks' Lambda | 0.4899746 | 2.5186 | 0.0006*** | 0.009** | 5.123 | 0.0005*** |
| | Pillai's Trace | 0.5939683 | 2.3019 | 0.0016** | 0.024* | | |
| **Inferior Longitudinal Fasciculus Right** | Wilks' Lambda | 0.4098067 | 3.3412 | 0.00001*** | 0.00015*** | 5.5868 | 0.0002*** |
| | Pillai's Trace | 0.75312 | 3.1545 | 0.00001*** | 0.00015*** | | |
| **Superior Longitudinal Fasciculus II Left** | Wilks' Lambda | 0.2363805 | 5.4462 | 0.00001*** | 0.00015*** | 12.1586 | 0.00001*** |
| | Pillai's Trace | 1.0818574 | 4.6713 | 0.00001*** | 0.00015*** | | |
| **Superior Longitudinal Fasciculus II Right** | Wilks' Lambda | 0.3109093 | 4.0814 | 0.00001*** | 0.00015*** | 10.7296 | 0.00001*** |
| | Pillai's Trace | 0.8199872 | 3.1459 | 0.00001*** | 0.00015*** | | |
| **Uncinate Fasciculus Left** | Wilks' Lambda | 0.5185115 | 2.4073 | 0.001** | 0.015* | 4.3313 | 0.0018** |
| | Pillai's Trace | 0.5730366 | 2.3076 | 0.0015** | 0.0225* | | |
| **Uncinate Fasciculus Right** | Wilks' Lambda | 0.3974788 | 3.6319 | 0.00001*** | 0.00015*** | 6.5693 | 0.00001*** |

| | | | | | |
|---|---|---|---|---|---|
| | Pillai's Trace | 0.780231 | 3.441 | 0.00001*** | 0.00015*** |

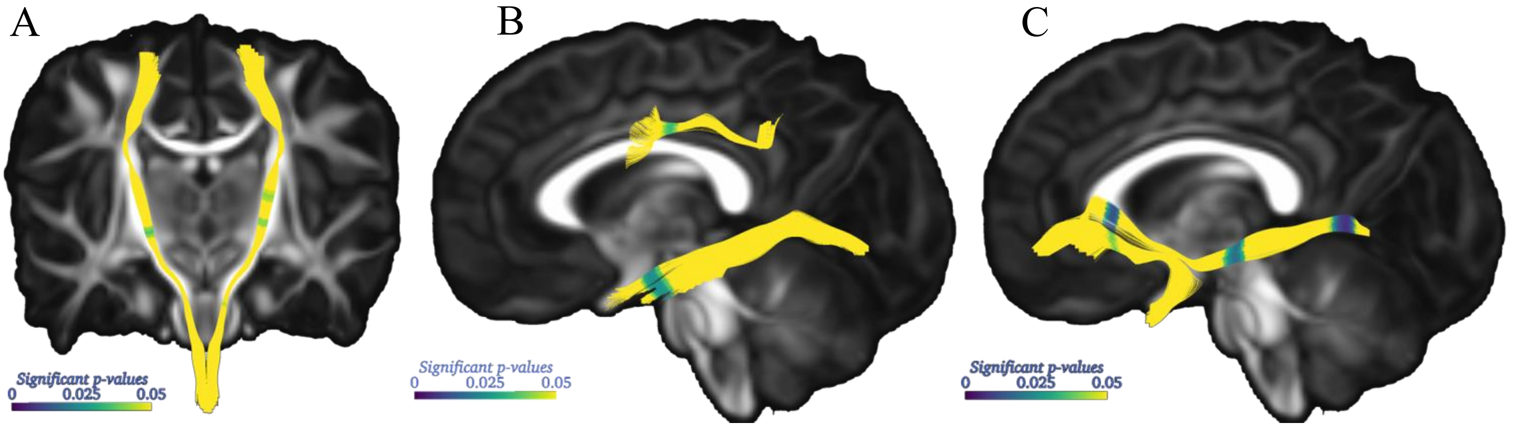

**eFigure 2. Visualization of the significant *p*-values along the tracts generated using Slicer.** (A) displays significant regions of RD in the bilateral CST, (B) highlights significant RD in the left SLF II and ILF, and (C) shows significant FA in the right IFOF and UNC. Abbreviations: IFOF, inferior fronto-occipital fasciculus; ILF, inferior longitudinal fasciculus; SLF, superior longitudinal fasciculus; CST, corticospinal tract; UNC, uncinate fasciculus. FA, fractional anisotropy; RD, radial diffusivity.
